# Neuromechanics of nonuse and compensation: implications for rehabilitation after stroke

**DOI:** 10.1101/2025.09.11.25335554

**Authors:** Germain Faity, Denis Mottet, Jérôme Froger, Marion Delorme

**Author notes:** Correspondence: Germain Faity; 11 avenue Robert Schuman, 35170 Bruz, France.

## Abstract

Stroke often results in compensatory movement strategies that combine use of trunk movements with nonuse of the affected upper extremity, which is considered detrimental to recovery. In this study, we investigate whether compensation and nonuse can be more than a suboptimal counterpart to recovery. We explore the influence of neuromechanics on nonuse and compensation in 22 stroke survivors and 22 controls. We asked seated participants to reach a target under two nonuse conditions (spontaneous or with minimized trunk compensation) and under three arm weight conditions (control, lightened and weighted). We replicated the findings that stroke induces more trunk compensation, more anterior deltoid activation and greater effort. We found that, in stroke patients, reduced arm weight decreased nonuse, while in controls, increased arm weight induced trunk compensation similar to what is observed after stroke. Furthermore, reaching with nonuse after stroke required less effort and preserved a force reserve at the shoulder. Our results show that both poststroke and healthy individuals engage in similar nonuse and compensation in the face of their neuromechanical limitations, as predicted by an optimal control policy aiming to succeed at the task while keeping a force reserve at each joint. We derive two theoretical predictions for future clinical research. First, patients whose nonuse yields few functional gains should benefit most from interventions to reduce compensation. Second, improving shoulder strength after stroke should reduce relative muscle activation, thereby decreasing trunk compensation and upper extremity nonuse. Clinical Trial: NCT04747587

## INTRODUCTION

Stroke often results in the nonuse of the affected upper limb, leading to a secondary functional decline, a phenomenon well-captured by the “use it or lose it” principle^1^. Although the learned nonuse theory has driven important advances in stroke rehabilitation^2^, emerging evidence suggests more nuanced underlying mechanisms.

The theory posits that pain, effort, and failure experienced during attempts to use the affected limb can discourage patients, prompting them to adopt compensatory strategies such as relying on their less affected arm. This avoidance behavior can persist even after improvement of arm function^2^, leading to non-mandatory compensations and nonuse of the paretic arm. Nonuse would thus stem from patients’ difficulties in accurately assessing the progression of their impairments and making informed decisions^3^.

Constraint-induced movement therapy (CIMT) is a therapeutic intervention based on the learned nonuse theory that has proven effective in promoting arm use^4^. Yet, the sustained arm use benefits may diminish over time, and the functional improvements do not always correlate with enhanced performance or ability^2,5,6^. Moreover, CIMT may encourage compensatory strategies, notably increased trunk involvement^7,8^. Interestingly, when instructed, many patients can reduce such trunk compensations^9^, suggesting that nonuse is not solely about limb preference but also influences broader movement patterns, as evidenced by the Proximal Arm NonUse (PANU) test^9^. Recent studies also indicate that many post-stroke patients are aware of their paretic arm’s potential^10,11^, challenging the hypothesis that nonuse stems solely from difficulties in accurately assessing impairments^3^. While the learned nonuse perspective aligns with prevailing views within the broader rehabilitation community, there remains a disparity between the theory and actual evidence^12^, calling for a broader understanding of nonuse mechanisms to improve patient selection and treatment allocation^13^.

An alternative perspective on nonuse emerges from the neuromechanical understanding of human movement. Neuromechanics is an integrative approach in which the behavior is an optimal response to interactions between neuronal, biomechanical and task constraints^14^. According to this approach, nonuse is possible because the human body offers an abundance of mechanical and neural solutions to meet the demand of the task, thereby allowing to choose the most efficient solution.

In a healthy individual, the central nervous system (CNS) orchestrates coordination strategies that optimally balance the competing objectives of maximizing success and minimizing effort^15^. Maximizing success means shaping movement from a set of task-related constraints, combining feedforward and feedback control. Minimizing effort means shaping movement from a set of efficiency rules combining mechanical and neural costs^16^. Notably, as the energy cost increases with muscle activation^17^, it makes sense to limit excessive muscle activations by minimizing the sum of squared motor command. This also reduces motor noise, which improves accuracy and controllability of movements^18^. This approach has elucidated the computational structure of the motor control system^19^, as well as anticipatory postural adjustments^20^.

Following a stroke, impaired neuromechanics redefine the concept of optimal functioning^21^. Lesions in the motor cortex and corticospinal tract disrupt the recruitment and synchronization of motor units, resulting in muscle weakness^22^—a primary contributor to post-stroke disabilities that significantly affects quality of life^23,24^. Compensatory strategies such as increased trunk movement or reliance on the less affected arm emerge due to saturated shoulder activation in simple reaching movements^25^. Thus, for some patients, compensatory strategies are necessary to perform tasks due to insufficient strength. For others, although they could perform the movement without compensation, they engage in trunk compensation. We believe that this non-mandatory compensation helps them avoid high muscle activations, thereby minimizing effort and optimizing arm function. This phenomenon mirrors observations in healthy volunteers, where increasing arm weight limits proximal joint use and induces trunk compensation^26^, which is the optimal solution to succeed at the task while minimizing antigravity muscle saturation^27^.

Therefore, while nonuse in some patients may be driven by operant conditioning, i.e. avoidance learning from adverse movement experiences that persists even after functional recovery (learned nonuse), it is also possible that nonuse in others is driven by a strategy of effort conservation (optimal nonuse).

In the present study, we investigate how effort minimization contributes to nonuse of the affected upper limb after stroke. We hypothesize that, in a seated reaching task, compensatory trunk movements will increase to minimize antigravity muscle saturation, both in healthy individuals and those with stroke. To test this hypothesis, we mimicked a deficit in shoulder strength for healthy participants using a heavy dumbbell and reduced the consequences of the deficit in stroke participants using an antigravity system. We then discuss the possibility of distinguishing between the two types of nonuse among patients and provide insights into tailored treatment approaches accordingly.

## METHODS

### 1. Participants

We enrolled 22 patients with stroke and 22 age– and sex-matched controls. Inclusion criteria for stroke participants were: first supratentorial stroke, at least one month post-stroke, and the ability to touch the opposite knee with the paretic arm while seated. Control participants had no history of stroke or upper limb impairments. Exclusion criteria included inability to provide informed consent, cognitive challenges precluding understanding of the study, or other conditions that could affect task performance.

All participants provided written consent. This study adhered to the 1964 Declaration of Helsinki and was approved by the Nimes Ethics Committee (N°ID-RCB: 2020-A02695-34, Clinical Trial: NCT04747587). To minimize bias, the specific study objective was not disclosed to participants.

### 2. Procedure

Participants completed the Proximal Arm Nonuse (PANU) test (Figure 1), comprising two tasks: a first reaching task without instructions (Spontaneous Arm Use, SAU) followed by a reaching task with instructions to minimize trunk movements (Maximal Arm Use, MAU)^9^. The distance to the target was set to match each participant’s arm length, determined from the medial axilla to the distal wrist crease. Participants performed 3 successive tests, from which we retained the median score.

**Figure 1.**
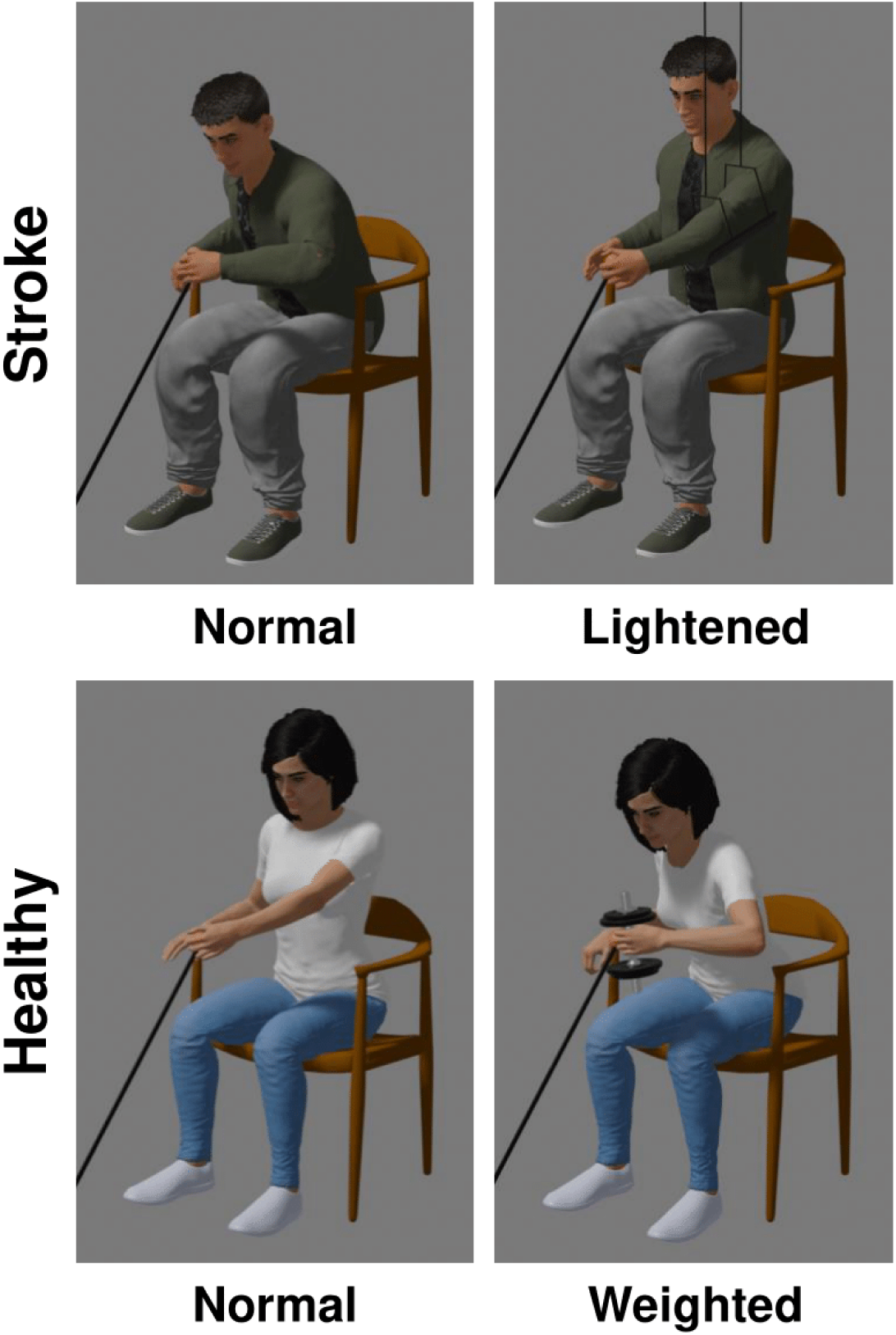
Experimental conditions for the nonuse and compensation assessment. Each panel represents the spontaneous arm use conditions of the PANU test, performed by participants with stroke (top row) or healthy participants (bottom row). Both groups performed the test with normal arm weight (left column). Participants with stroke also performed the test with a lightened arm (top right) and healthy people with a weighted arm (bottom right).

All participants performed the test with their inherent arm weight (control condition). Additionally, stroke patients completed the test with an antigravity system (lightened condition) and healthy participants executed the test while holding a dumbbell (weighted condition). Test conditions were randomized and interspersed with rest periods sufficient to mitigate fatigue. The dumbbell weight was selected to induce an antigravity effort of 85% of the shoulder’s maximal voluntary force^26^. The antigravity force resulted in a complete reduction in the weight of the arm.

**Table 1.**
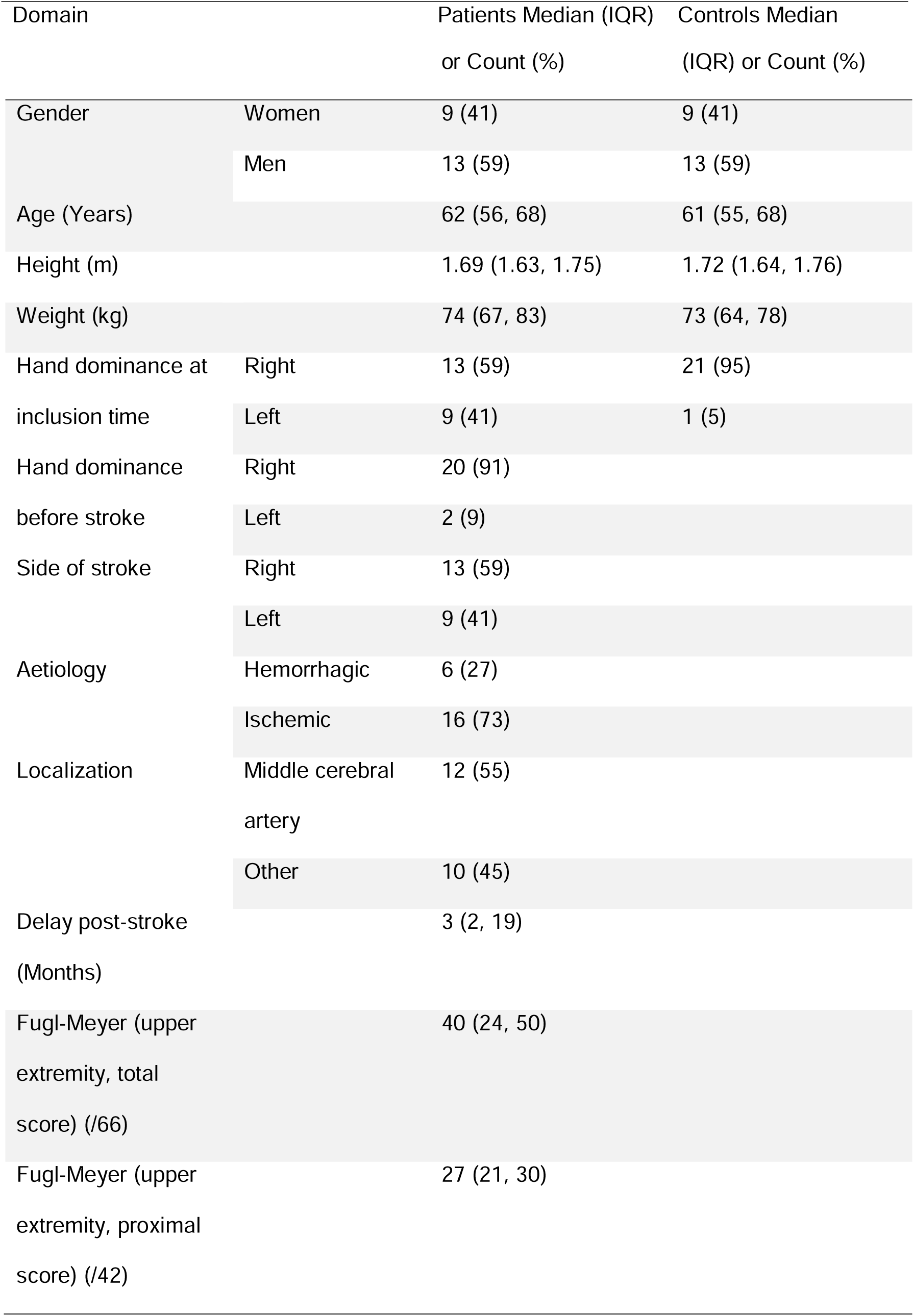
Demographic, anthropometric, and clinical characteristics of the post-stroke (N=22) and control (N=22) participants.

### 3. Experimental Setup

Movements were recorded at 30 Hz using the Kinect V2 markerless motion capture system (Microsoft, Redmond, WA, USA), which has been proven sufficiently accurate for this type of task^28,29^. The Kinect sensor was positioned in front of the participant at a distance of 1.50 meters.

Muscle activation was recorded at 1258 Hz with surface EMG (Delsys Trigno Wireless System, USA). We recorded six muscles involved in shoulder movements (anterior deltoid, medial deltoid, biceps brachii, clavicular head of pectoralis major) and trunk movements (right-sided longissimus, left-sided longissimus).

After each reaching task, participants rated their perceived exertion on a Borg scale ranging from 6 (no effort) to 20 (maximal effort)^30^. This scale accurately reflects actual effort in upper limb tasks^31^.

### 4. Data Analysis

Data analysis was performed using Scilab 6.0.2.

#### 4.1 Motion Capture

First, all position time series were low-pass filtered at 2.5 Hz using a double-pass second-order Butterworth filter. The 2.5 Hz frequency was chosen to retain 95% of the motion signal while eliminating high-velocity errors^29^.

Second, the beginning of the movement (t_0_) was defined as the point at which the Euclidean velocity of the hand in task space became positive and remained so until the maximum velocity was reached. The end of the movement (t_final_) was defined as the point at which the Euclidean distance to the target reached its minimum^9^.

Third, shoulder displacement was calculated as the Euclidean distance between the shoulder position at t_0_ and t_final_.

#### 4.2 Torque

Antigravity shoulder torque was calculated as the Euclidean norm of the 3D static torque against gravity at t_final_, divided by the participant’s maximum shoulder torque, to provide a percentage of the maximal antigravity torque at the shoulder. To calculate the final posture static torque, the positions of the center of mass and the absolute weight of the upper limbs were approximated using De Leva’s equations^32^, based on the participant’s height and weight.

#### 4.3 Muscle Activation

All EMG time series were bandpass filtered between 10 and 850 Hz using a double-pass second-order Butterworth filter (80dB/dec). The root mean square (RMS) envelope was obtained by applying a moving average to the absolute value of the EMG signal with a 50 ms epoch. For each muscle, EMG was normalized by the EMG at maximum voluntary contraction (MVC) to quantify muscle saturation (i.e., % muscle use). For each reach and each muscle, muscle activation at the final posture was calculated as the mean normalized EMG when the hand was close to the target (hand-to-target distance <5% of the reach length).

### 5. Statistical Analysis

All statistical analyses were conducted using R 4.2.1 with the rstatix package (version 0.7.1). Normality was verified using quantile-quantile plots. For trunk compensations, anterior deltoid activation, antigravity shoulder torque, and rating of perceived exertion data, ANOVAs were conducted with group (between factor: stroke versus healthy), arm use (within factor: spontaneous versus maximal), and arm weight (within factor: light versus heavy) as factors. Significance was set at p < .05. Effect sizes are reported using partial eta squared (η²_p_). Data are presented as mean (± standard deviation).

## RESULTS

### 1. Shoulder Maximum Voluntary Torque

A two-way ANOVA revealed a greater difference in shoulder antigravity torque between paretic and non-paretic arms in stroke patients compared to the difference between non-dominant and dominant arms in healthy individuals (F(1,42) = 16.656, p < .001, η²_p_ = .28, stroke difference = 46%, healthy difference = 9%) (Figure 2).

**Figure 2.**
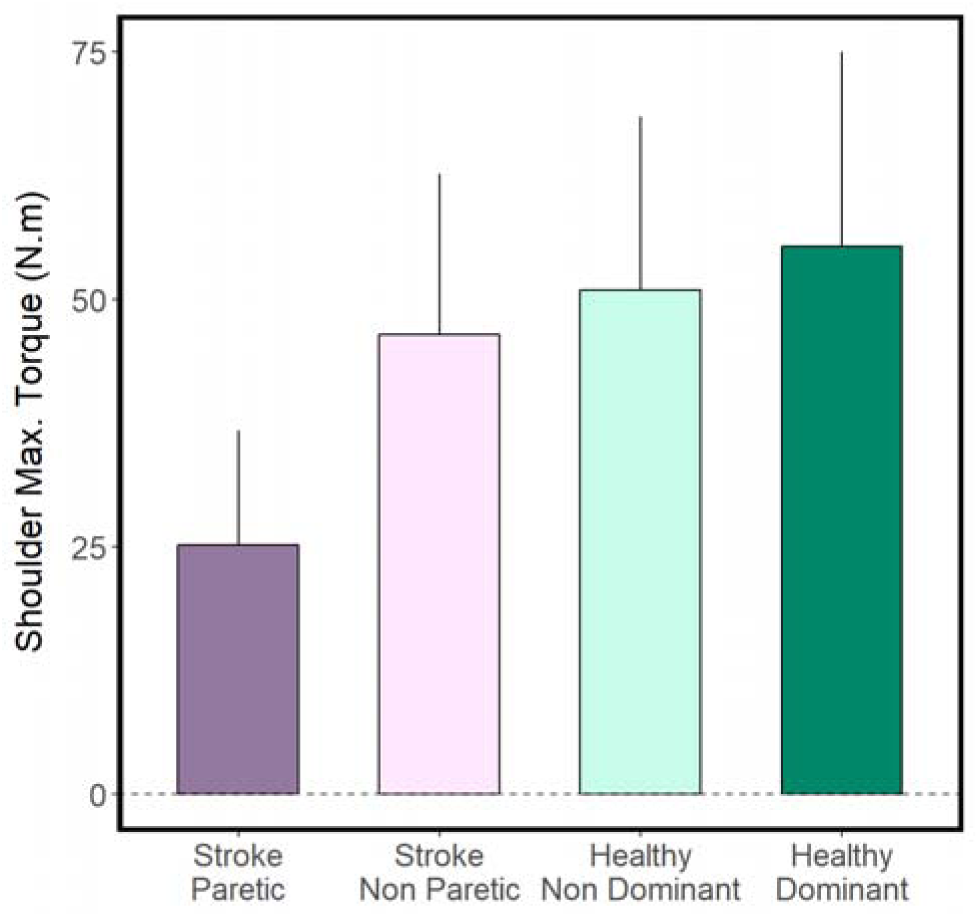
Maximum Voluntary Antigravity Torque at the Shoulder for both upper limbs in individuals with or without stroke. The difference in shoulder torque between the paretic and non-paretic arms is greater than the difference between non-dominant and dominant arms in healthy participants. Data are presented as mean (± standard deviation over participants).

### 2. H1: Trunk Compensations Modulate The Effort Required In Both Stroke And Healthy Participants When Reaching

#### Trunk Compensation

A two-way ANOVA (F(1, 40) = 20.208, p < .001, η²_p_ = .34) showed stroke participants used more trunk compensations in the spontaneous condition than in the maximal condition (F(1, 18) = 37.800, p < .001, η²_p_ = .68), while no significant difference was found in healthy participants (F(1, 22) = 4.020, p = .057, η²_p_ = .16) (Figure 3A).

**Figure 3.**
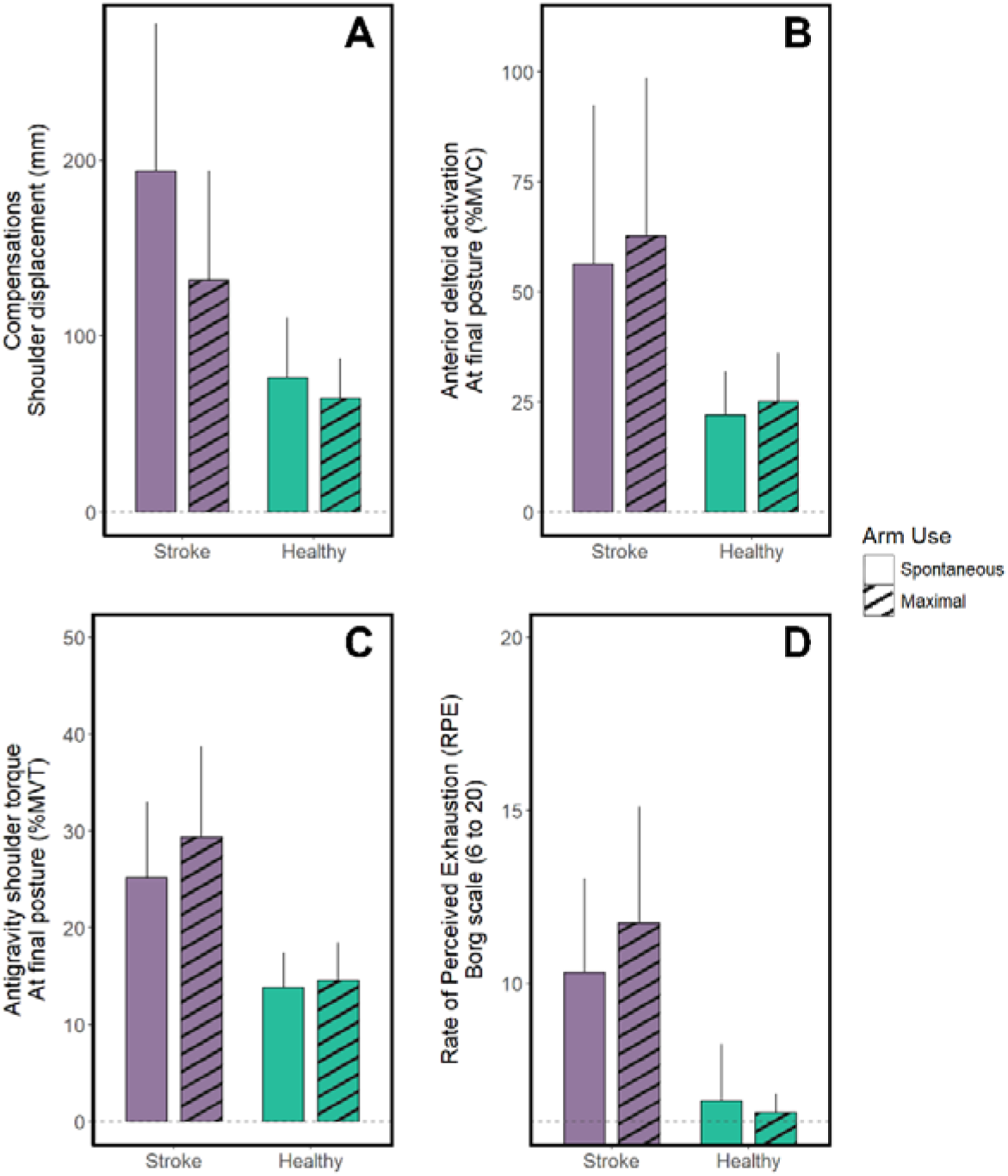
Effects of Spontaneous vs. Maximal Arm Use in individuals with or without stroke. Panel A shows the increased reliance on trunk compensation after stroke. Panel B and C show similar effects, with an increased deltoid activation and an increased antigravity shoulder torque during maximal arm use, more pronounced after stroke. Panel D shows the higher perceived exertion in the maximal condition after stroke, but not in healthy participants. All data are presented as mean (± standard deviation over participants).

#### Anterior and Medial Deltoid Activation

A two-way ANOVA revealed higher anterior deltoid activation in stroke patients in both conditions (F(1, 40) = 21.621, p < .001, η²_p_ = .35). Both groups increased deltoid activation in the maximal condition compared to the spontaneous condition (F(1, 40) = 9.308, p = .004, η²_p_ = .19) (Figure 3B). Similar effects were observed for the medial deltoid (Stroke vs Healthy: F(1, 40) = 28.186, p < .001, η²_p_ = .41; Spontaneous vs Maximal: F(1, 40) = 5.122, p = .029, η²_p_ = .11).

#### Antigravity Shoulder Torque

A two-way ANOVA revealed that the increase in antigravity shoulder torque in the final posture was greater after stroke than in healthy individuals (F(1, 40) = 12.851, p = .001, η²_p_ = .24).

#### Rate of Perceived Exertion

A two-way ANOVA (F(1, 40) = 8.464, p = .006, η²_p_ = .18) revealed that stroke participants perceived greater exertion in the maximal condition than in the spontaneous condition (F(1, 18) = 5.720, p = .028, η²_p_ = .24). No significant difference was found in healthy participants (F(1, 22) = 1.840, p = .188, η²_p_ = .08) (Figure 3D).

This first set of analyses demonstrates that the reaching task is more resource-intensive for stroke participants. When reaching spontaneously, stroke participants compensated with the trunk more than necessary (nonuse), which allowed them to reduce the required effort.

### 3. H2: Arm Weight Modulates Nonuse, Compensations, and the Effort Required in Both Healthy and Stroke Participants

#### Trunk Compensation

Reducing arm weight decreased trunk compensation by 15% in the spontaneous condition for stroke patients but not in the maximal condition (Stroke arm weight X arm use: F(1, 13) = 4.81, p < .001, η²_p_ = .27; Spontaneous: F(1, 13) = 18.6, p < .001, η²_p_ = .59; Maximal: F(1, 13) = 1.62, p = .225, η²_p_ = .11). In healthy participants, increasing arm weight resulted in a 106% increase in trunk compensation in the spontaneous condition and 90% in the maximal condition (Healthy arm weight X arm use: F(1, 18) = 11.536, p = .003, η²_p_ = .39; Spontaneous: F(1, 18) = 54.3, p < .001, η²_p_ = .75; Maximal: F(1, 18) = 22.0, p < .001, η²_p_ = .55) (Figure 4A).

**Figure 4.**
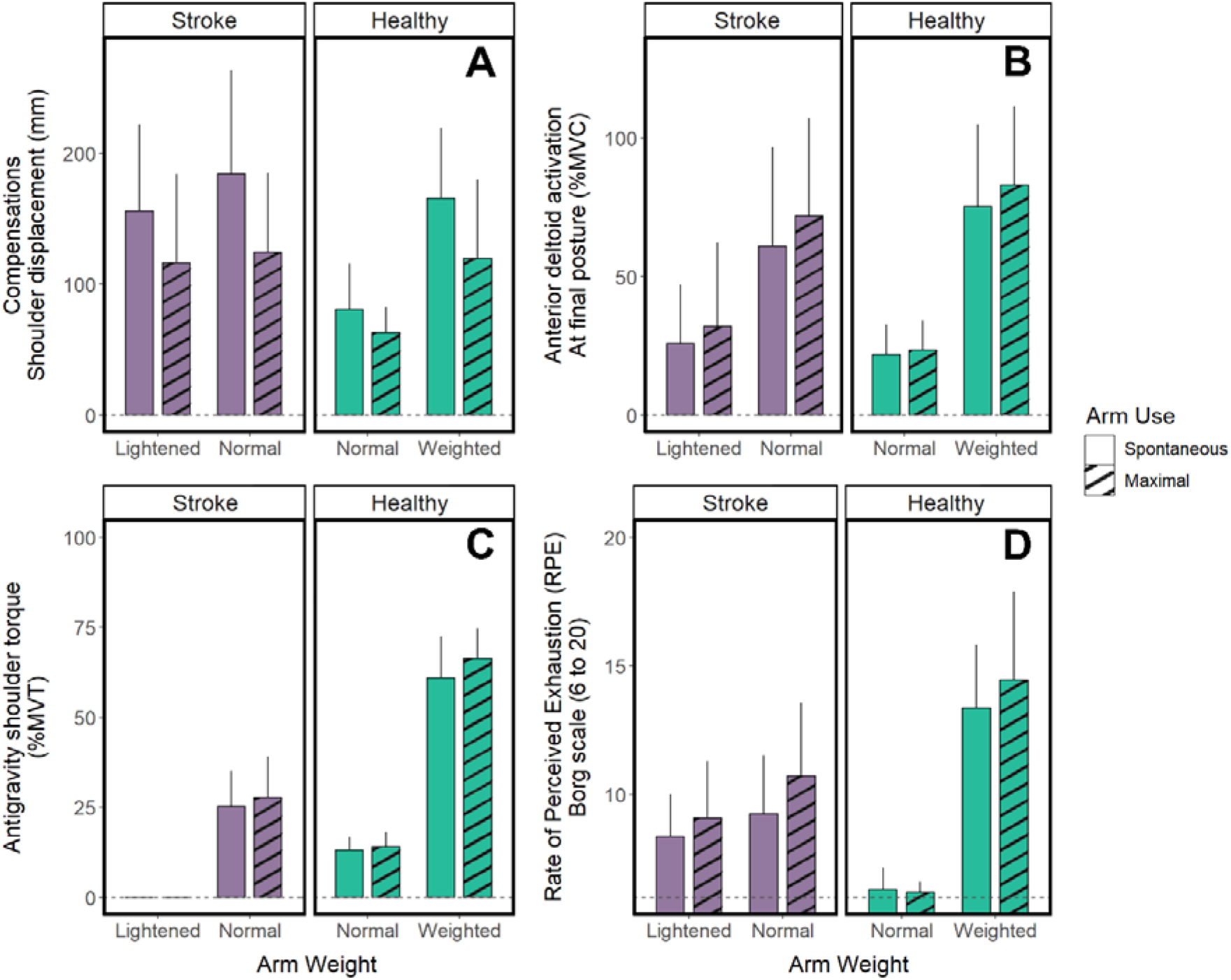
Effect of stroke, arm weight and arm use on Trunk Compensation, Anterior Deltoid Activation, Antigravity Shoulder Torque, and Perceived Exertion. The figure shows that, in people with and without stroke and in spontaneous and maximal conditions, trunk compensation increases with arm weight (A), anterior deltoid activation increases with arm weight (B), antigravity shoulder torque increases with arm weight (C) and perceived exertion increases with arm weight (D). Data are presented as mean (± standard deviation over participants).

#### Anterior and Medial Deltoid Activation

Reducing arm weight decreased anterior deltoid activation by 56% in stroke patients in both spontaneous and maximal conditions (Stroke arm weight X arm use: F(1, 13) = 0.696, p = .419, η²_p_ = .05; arm weight main effect: F(1, 13) = 80.2, p < .001, η²_p_ = .86). Increasing arm weight increased anterior deltoid activation by 250% in healthy participants in both spontaneous and maximal conditions (Healthy arm weight X arm use: F(1, 18) = 3.827, p = .07, η²_p_ = .18; arm weight main effect: F(1, 18) = 106.722, p < .001, η²_p_ = .86) (Figure 4B). Similar effects were observed for the medial deltoid (Stroke: F(1, 13) = 23.708, p < .001, η²_p_ = .65; Healthy: F(1, 18) = 82.458, p < .001, η²_p_ = .82).

#### Antigravity Shoulder Torque

Increasing arm weight increased antigravity shoulder torque in healthy participants in both conditions (Healthy arm weight X arm use: F(1, 18) = 6.806, p = .018, η²_p_ = .27; Spontaneous: F(1, 18) = 434, p < .001, η²_p_ = .96; Maximal: F(1, 18) = 715, p < .001, η²_p_ = .98). Both groups increased torque in the maximal condition compared to the spontaneous condition, regardless of arm weight (Stroke: F(1, 12) = 7.436, p = .017, η²_p_ = .36; Healthy control arm weight: F(1, 18) = 7.79, p = .012, η²_p_ = .30; Healthy loaded arm weight: F(1, 18) = 9.85, p = .006, η²_p_ = .35) (Figure 4C).

#### Rate of Perceived Exertion

Reducing arm weight decreased perceived exertion by 13% in stroke patients in both the spontaneous and maximal conditions (Stroke arm weight X arm use: F(1, 13) = 2.567, p = .133, η²_p_ = .17; arm weight main effect: F(1, 13) = 7.612, p = .016, η²_p_ = .37). Increasing arm weight increased perceived exertion by 112% in the spontaneous condition and 133% in the maximal condition for healthy participants (Healthy arm weight X arm use: F(1, 18) = 5.570, p = .030, η²_p_ = .24; Spontaneous: F(1, 18) = 157, p < .001, η²_p_ = .90; Maximal: F(1, 18) = 107, p < .001, η²_p_ = .86) (Figure 4D).

This second set of analyses demonstrates that decreasing arm weight in participants with stroke reduces both compensations and the effort required. Conversely, in healthy participants, increasing arm weight leads to compensations similar to those observed after stroke. Thus, when their arm is weighted, healthy participants exhibit nonuse, enabling them to reduce the required effort.

## DISCUSSION

In this study, we leverage principles from neuromechanics and optimal control theory to better understand the reasons for nonuse of the affected upper limb after stroke. These principles predict that, in a seated reaching task, arm nonuse combined with trunk compensation is an optimal strategy to reduce antigravity effort at the shoulder when arm weight is substantial compared to the available strength.

Our findings indicate that the nonuse of the paretic arm, facilitated by trunk compensation, reduces the effort required at the shoulder to maintain the arm near the target in both participant groups. These compensations, frequently observed in post-stroke patients, also emerge spontaneously in age-matched healthy subjects when the arm’s weight is substantial compared to their available strength. However, while compensations and effort decrease in individuals with stroke when arm weight is experimentally reduced, they never reach the baseline levels observed in healthy participants.

We discuss below how and why the strategy of nonuse and compensation emerges as an optimal solution under task and participant constraints. We translate this theoretical insight into a clinical vision where treatments are individualized based on neuromechanics. Patients with enough remaining function who can voluntarily cancel compensations without loss of function or increased effort should benefit most from therapies aimed at minimizing compensations. Conversely, those who cannot cancel compensations without experiencing loss of function or increased effort should benefit from therapies to improve shoulder strength.

### 1. Compensation and Nonuse Emerge to Save Neuromechanical Effort

We found that, in both healthy individuals and stroke survivors, trunk compensation and nonuse of the upper limb emerge when the force-to-weight ratio approaches the neuromechanical limits. When we experimentally increased arm weight in healthy individuals, this imposed a higher neuromechanical effort at the shoulder (higher antigravity activations Figure 4B, higher antigravity torque Figure 4C and higher perceived effort Figure 4D). Faced with such constraints, individuals modified their movement organization, particularly visible with the doubling of trunk displacement in healthy individuals (Figure 4A). This pattern mirrors the movement and nonuse consequences observed in stroke participants when the arm is not lightened (Figure 3).

How do we account for such behavioral adaptation in both healthy people and stroke victims? We believe this is mainly because we optimally organize our movements to maximize reward and minimize effort^33^. Maximizing reward means succeeding in the reaching task, i.e. zeroing out the hand-target distance. Minimizing effort means selecting movements with low foreseeable costs, i.e. avoiding high motor commands. In sum, we adopt an optimal feedback control policy to form the successful hand trajectory by moving the trunk-arm-forearm segments while investing the least effort possible^15,16,34,35^. Mathematically, this is tantamount to minimizing the sum of squared motor commands, which captures ATP consumption in muscles^16^ and also leads to the distribution of work across multiple joints^19^. A minimal neuromechanical model of the seated reaching task revealed that proximal arm nonuse combined with trunk compensation is the optimal solution for reducing shoulder antigravity effort: when arm weight exceeds a certain proportion of the available shoulder force, a novel movement organization becomes optimal because compensation with the trunk distributes the exertion and reduces shoulder workload^27^.

Following this logic, we hypothesized that reducing arm weight would subsequently lower the required percentage of shoulder muscle activation for the task^36^, leading to diminished trunk compensations. Our data corroborate this hypothesis: a decrease in arm weight causes reduced compensations (Figure 4A), decreased shoulder activation (Figures 4B and 4C), lessened perceived exertion (Figure 4D), and ultimately, enhanced arm use. Our results are predicted by optimal control theory and corroborate previous studies^31,37–39^, underscoring the central role of compensation and nonuse in limiting effort when arm weight exceeds a significant proportion of available shoulder force.

Consequently, the widespread idea that nonuse with compensation is a suboptimal counterpart to recovery is too superficial a vision^38,40^. Here, we provide evidence that at least a portion of the nonuse emerges from an optimal sensorimotor organization within a deficient human body.

### 2. The Role of Sensorimotor Learning

Even with a lightened arm limiting the strength deficit, participants with stroke exhibited residual compensatory behaviors, surpassing the trunk recruitment seen in healthy controls (Figure 4A). This observation underscores that the effects of stroke extend beyond strength deficits, disrupting muscle coordination and hindering arm control. Although muscle activation patterns with a lightened arm resembled those of healthy participants, our data suggest that nonuse motor strategies might serve dual purposes: reducing effort and enhancing arm control. This control improvement might be aided by a greater reliance on the trunk, which is less impacted by the effects of a stroke^41^. Additionally, discomfort or pain from moving the paretic arm could discourage certain movements, aligning with optimization principles. Sensory impairments or spasticity may also contribute to nonuse under the same principles. However, evidence suggests that their influence is less pronounced than deficits in strength, which appear to be the main predictor of reaching performance after stroke^42^.

Conversely, unfamiliarity with the arm-lightening apparatus might contribute to the residual compensations observed, akin to behaviors seen under microgravity conditions^43^. Extensive training could potentially counteract these compensatory actions^44^.

The learned nonuse theory also suggests that movement choices often represent local minima in motor control, i.e. strategies that approach but do not achieve global optimization. These can be modified through coordination adjustments. Such local minima have been observed in healthy individuals^45^. The central nervous system may often target these local solutions as the first operational strategy to succeed at the task, before broader exploration reveals even more effective movement strategies. This exploration might arise from movement variability, observing skilled performers, or external constraints. Recent research indicates that even after experiencing optimal motor strategies, healthy people frequently revert to familiar movement patterns, reflecting a kind of ‘motor memory’ that can impede new learning^48,49^. A similar scenario is seen in sports, like high jumpers transitioning between the scissors jump and the Fosbury flop. Although these transitions might be initially challenging, they can ultimately lead to superior performance, paralleling the concept of learned nonuse.

### 3. Clinical Implications

Immediately following a stroke, nonuse is likely the best strategy, because although various strategies are possible, compensation conserves effort while maintaining improved function. However, after a period of spontaneous recovery and rehabilitation, during which the function of the paretic arm improves, nonuse may remain the best strategy for some individuals, while for others, it may become a learned strategy that is no longer optimal. Historically, Taub^2^ described this phenomenon as “learned nonuse”, attributing it primarily to operant conditioning, but the term is today often used by clinicians to refer to all forms of nonuse^12^. Our findings suggest a more nuanced understanding: learned nonuse as defined by Taub should no longer be confused with what we here refer to as “optimal nonuse”.

Here we show that optimal nonuse is driven by effort conservation, whereas learned nonuse is driven by operant conditioning^2^. This distinction has clinical implications.

#### A Method to Identify Participants Most Likely to Benefit from Therapies Aimed at Minimizing Compensations

Building on our understanding of these two types of nonuse, we explore methods to identify which stroke survivors would benefit most from therapies aimed at minimizing compensations, such as CIMT^2,4^ or trunk-restraint^47^. Our data serves as a foundational example (Figure 5).

**Figure 5.**
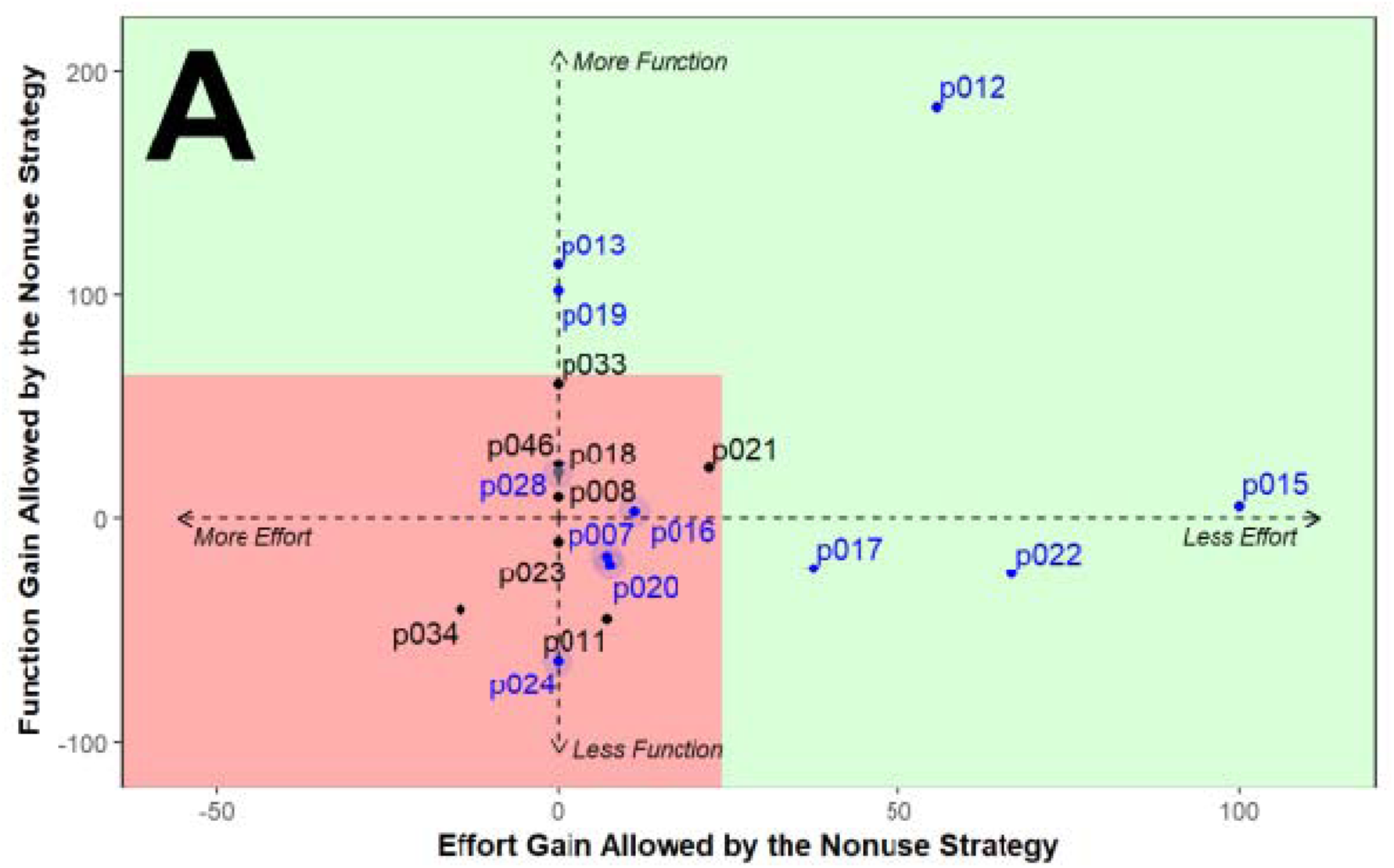
Gains from nonuse plus compensation strategies. The gain in effort (horizontal axis) is assessed from the rate of perceived exertion (RPE). The gain in function (vertical axis) is assessed from the movement time (MT). Gains are quantified by the difference (in RPE or MT) between the MAU and SAU conditions of the PANU test and expressed as a percentage. Black dots indicate patients without nonuse, and blue dots indicate patients with nonuse. The red zone is the region that encloses all black dots, i.e. the area where nonuse is not beneficial. Outside this region, the green zone denotes cases where nonuse is beneficial (for effort or function). Among blue dots, patients can either show no benefit (they fall in the red zone) or varied benefit from their nonuse (they fall in the green zone; e.g., p012 gains both function and effort, p013 & p019 gain function, p017 & p022 gain effort). The clinical implications of this figure are as follows. Because they gain little with nonuse plus compensations, patients in the red zone may benefit most from targeted interventions aimed at reducing trunk compensations. Conversely, because they gain with nonuse plus compensations, patients in the green zone could benefit most from other interventions, such as those aimed at increasing shoulder strength.

According to the neuromechanical framework, patients who demonstrate improved function or reduced effort from nonuse strategies do so because it is the optimal solution for them. Preventing these compensations will either increase the effort required to perform movements or decrease arm function. In Figure 5A, such patients include p012, p013, p019, p015, p017, and p022 and can be considered to have “optimal nonuse”. In contrast, patients such as p007, p016, p020, p024, and p028 do not show significant gains from the nonuse strategy, which is more consistent with “learned nonuse”.

By distinguishing between learned nonuse and optimal nonuse, therapists can tailor treatments more effectively. For patients with learned nonuse, therapies aimed at reducing nonuse, such as CIMT or TRMT, may be most beneficial. These therapies can help these patients overcome motor memory barriers and foster further progress. Conversely, patients with optimal nonuse, may benefit more from interventions aimed at increasing shoulder strength to counteract the deficits affecting arm function or effort. This distinction should not be interpreted as strictly binary. Nonuse is likely to fall along a continuum, with some patients showing mixed profiles depending on task demands. Nonetheless, this framework provides a valuable basis for helping therapists identify the most appropriate therapies for their patients. In summary, conducting a PANU test using a low-cost markerless motion capture system (Kinect v2)^29^ allows therapists to better tailor treatments, potentially enhancing rehabilitation outcomes.

While our selection process is theoretically sound, it is limited by the reliability and generalizability of the results. Different tasks may yield varying outcomes, highlighting the importance of comprehensive multi-task and multi-day analyses to fully understand how compensations affect function and effort for each patient^48^. Future studies should aim to determine the effectiveness of this proposed methodology in identifying patients who would benefit most from interventions aimed at reducing compensatory behaviors.

#### The Role of Strength Training

We found that increasing arm weight or decreasing shoulder strength increase muscle activation (Figure 4). The mirror image of these results is that either decreasing arm weight or increasing shoulder strength should reduce muscle activation to a level accessible by stroke patients, providing a sound rationale for therapeutic interventions. The benefits of decreasing arm weight is well known to facilitate repetitive task training, especially with robotic devices^49,50^. The benefits of strength training are also suspected; clinical trials should explore customized shoulder and arm strength training for stroke patients^51^.

Beyond influencing peripheral factors such as hypertrophy, resistance training is a well-established method for increasing muscle activity in healthy individuals. It enhances motor unit recruitment, motor units coordination, and intermuscular coordination^52^. The neural mechanisms underlying these improvements are not fully understood; however, they may involve increased cortical excitability, facilitating more efficient transmission of motor commands along the corticospinal tract. Such improvements can further enhance force production and facilitate motor learning, both of which are critical post-stroke^53^.

Although there is evidence that resistance training can improve motor performance and even reduce spasticity in patients with stroke^54–56^, its integration into post-stroke rehabilitation is not yet consistent. This inconsistency may be due to a lack of conclusive evidence^51^ and potential deviations from established best practices^57^. However, incorporating insights from physical training into rehabilitation could optimize outcomes, possibly in synergy with intensive therapeutic approaches like CIMT. The primary aim would be to determine whether resistance training can enhance strength, thereby promoting improved arm use following a stroke.

### 4. Limitations

Our study specifically examined patients with moderate impairments (mean Functional Mobility Upper Extremity [FMUE] = 40, 95% Confidence Interval [CI] = [24, 50]). Consequently, our findings may not be directly applicable to those with severe impairments. For this latter group, merely enhancing shoulder strength through training may not suffice^58^. In such cases, employing exoskeletons or passive lightening systems to facilitate arm use could be more beneficial^59^.

Although our research focused primarily on seated reaches, previous studies have demonstrated a relationship between trunk compensations and arm use in daily activities^60^. This suggests that our findings might have broader implications for real-world applications.

## CONCLUSION

Our findings reveal that both stroke-affected and healthy individuals adopt nonuse of the paretic arm and trunk compensation when pushed towards their force limits. Drawing on neuromechanics and optimal control theory, we derive two predictions for better individualizing treatment allocation after stroke. First, patients who gain little functional and effort benefits from the nonuse strategy should benefit most from interventions aimed at reducing compensations. Second, for patients who can voluntarily limit compensations, improving shoulder strength should reduce relative muscle activation, decrease compensatory behaviors, and enhance arm use. Future research is needed to validate our theoretical predictions and their relevance to therapeutic decisions, as well as to better identify the most effective rehabilitation strategies.

## ACKNOWLEDGMENTS

This study was supported by Nîmes University Hospital (N°ID-RCB: 2020-A02695-34) and the LabEx NUMEV (ANR-10-LABX-0020) within the I-SITE MUSE. We would like to thank Simon Pla and Hugo Rodriguez for helping set up the experiment.

## DATA AVAILABILITY STATEMENT

The code and the datasets generated and analyzed during the current study are available in the Open Science Framework repository, https://osf.io/4mr7k/.

## AUTHOR CONTRIBUTIONS

Conceptualization, G.F., D.M., and J.F.; Methodology, G.F., D.M., and J.F.; Software, G.F. and D.M.; Validation, G.F., D.M., J.F., and M.D.; Formal Analysis, G.F., and D.M.; Investigation, G.F.; Resources, D.M., J.F., and M.D.; Data Curation, G.F.; Writing—Original Draft Preparation, G.F.; Writing—Review and Editing, G.F., D.M., J.F., and M.D.; Visualization, G.F., and D.M.; Supervision, D.M. and J.F.; Project Administration, G.F., D.M., J.F., and M.D.; Funding Acquisition, G.F., D.M. and J.F. All authors have read and agreed to the published version of the manuscript.

